# How was the Mental Health of Colombian people on March during Pandemics Covid19?

**DOI:** 10.1101/2020.07.02.20145425

**Authors:** Josimar E. Chire Saire

## Abstract

Actual pandemic started with first cases in China and fast spread in Europe, Asia and the next continents became a global concern from March, so the most of countries ordered a lockdown to decrease infection rate forced people to stay at home. The first confirmed case was on March 6, during the next days more cases were confirmed by Minster of Health, the first death was on March 21, one day after of the announcement of lockdown in Colombia. During this period of waiting for what is going to happen people can develop mental stress as fear, sleep disorders so to analyze how was the behaviour on the population the present study is conducted using a Social Network as Twitter, Natural Language algorithms and Data Mining criterion’s. The findings confirmed the interest of covid19 was increasing daily, people was experimenting fear, anxiety, mental stress.

## 1 Introduction

Covid19 is a pandemic which started on December 2019 in the city of Wuhan, China. This coronavirus was transmitted to many countries, one hypothesis about the fast growth was the actual global interconnection as result of globalization. World Health Organization declared the outbreak as global emergency on March 11, 2020^1^. Health Ministry confirmed the first case in Colombia on March 6^2^, this person arrived from Milan, Italy and had symptoms then went for health center for the analysis and National Health Institute confirmed as positive case. Two important points coming from official website of Ministry of Health: a) during the week of positive confirmed case, the minister had a meeting with Public, Private Health Staff to stablish a Response Plan, b) National Emergency Committee started 8 weeks before of the first case his work to promote self-care based in washing hands to the citizens. So, preparation phase ended and started mitigation phase in Colombian territory.

The evolution of cases after March 6^2^, two cases on March 9^3^ confirmed cases through a letter from Oficial Ministry of Health Twitter account, one man (34 years) from Valle del Cauca and a woman (50 years) from Medellin both coming from Spain. On March 11^4,5^, 4 new cases was reported in Medellin a close circle with the woman who came from Spain, 2 new cases in Bogota and one more in Cartagena, a total of 9 cases in Colombia. On March 12^6^, the president of Colombia, Ivan Duque announced covid19 as a national emergency so public events with more than 500 people was suspended and docking of ships and cruisers were prohibited.

Next day, March 13^7,8^ were reported 2 new cases in Bogota and two new in Neiva, two siblings living in Italy. Same day, three new cases^9^ were confirmed in Meta, Valle del Cauca, Bogota, and two came from Spain, the other one from Italy. Next day, March 14 there were 6 new cases^10^ in Bogota(2), Medellin(1) and Rionegro(3). Besides, the president on March 14^11^ announced the close of borders with Venezuela and strict control with Ecuador. On March 15^12^, the number grew to 10 new people Bogota(2), Cartagena(2), Cali(1), Cucuta(1), Dosquebradas(1), Manizales(1), Neiva(1) and Medellin(1), some of them were coming from Spain and one from Risaralda from United States of America(USA).

Later, new 11 cases^13^ were confirmed in Bogota(6), Neiva(4), Facatativa(1) and previously many of them travelled to Spain and one to USA. This day^14^, Ivan Duque, Colombia’s president ordered to suspend classes in schools and universities to decrease the curve of infections. On March 16^15^, 9 cases in Bogota and five of them travelled to Spain and one to Ecuador and the same day^16^, three more three in Bogota, previously two had a travel to Spain. On Tuesday^17^, Health Minister announced eight more cases in Cartagena(1), Barranquilla(2), Bucaramanga(1), Cali(2), Bogota(2) and many of them came from Spain(5), Italy(1), France(1). Hours after of this eight cases, 10 cases more were reported^18^ by the Minister in Bogota(7), Cucuta(2) and Cartagena(1) and recently travelled to Spain(3), USA(1), Turkey and Greece(1).

On March 18, Ministry of Healh announced^19^ new 18 cases in Armenia (2), Palmira (3), Cali (5), Pereira (1), Cajica (2), Tolima (1), Santander (1), Neiva (1) and Bogota (2). Same day, 9 cases^20^ more were reported in Bogota (3), Cartagena (2), Ibague (1), Pereira (2) and Dosquebradas (1), from this day no more details about where the people travelled or from which countries this people arrived. Next day(March 19) new 6 cases^21^ in Cartagena (1), Barranquilla (1), Palmira (1) and Medellin (3). This Friday(March 20), after midnight were confirmed 20 new cases^22^ in Bogotá (8), Cúcuta (3), Armenia (1), Popayán (2), Caldas (1), Neiva (1), Cartagena (1), Barranquilla (2) y Soacha (1) Health Ministry confirmed 17 cases^23^ in Pereira (1), Anapoima (1), Medellin (9), Cali (1), Rionegro (1), Bogota (3) and Envigado (1). This day, more 13 new cases^24^ were reported in Bogota (9), Santa Marta (2), Chia (1) and Madrid (1). Considering the previous situation, Ivan Duque as president of Colombia declared a national quarantine^25^ during the evening of March 20 and until this date, a total of 158 cases were reported.

Infodemiology^26^ is a new research field, with the objective of monitoring public health^27^ and support public policies based on electronic sources, i.e. Internet. Usually this data is open, textual and with no structure and comes from blogs, social networks and websites, all this data is analysed in real time. And Infoveillance is related to applications for surveillance proposals, i.e. monitor H1N1 pandemic with data source from Twitter^28^, monitor Dengue in Brazil^29^, monitor covid19 symptoms in Bogota, Colombia^30^. Besides, Social sensors is related to observe what people is doing to monitor the environment of citizens living in one city, state or country. And the connection to Internet, the access to Social Networks is open and with low control, people can share false information(fake news)^31^.

Cohen^32^ presents a graphic about the evolution of the pandemic during the first 100 days in Asia, Europe and North America and many of this countries would close their frontiers and require social distancing with costs to the Economy, Social Life, and Mental Health. Actually, there is a few studies about Covid19 Impact on Society, Bodrud^33^ conducted a psychosocial and socioeconomic study in Bangladesh based on the perception of the citizenship with 1066 participants and the findings were: there is no confidence to the governments action to deal with pandemic because of a weak health system, citizens have fear to loose one familiar for the lack of attendance so people decides to stay at home but this has implications producing mental and economic stress. Besides, a worrying about have problem to continue with education and socio-economic impact. Ahorsu^34^ published an analysis of many psychometric questionnaires to find a better for covid19 and the study was over 717 Iranian participants, and purposed a seven level scale of fear for covid19.

So, considering the pandemic and increasing number daily in Colombia, the proposal of this paper is analyzed the mental health of people living in Bogota, Colombia during the first month, March using social sensors based on publications, posts of citizenship to analyze how was the effect or impact of covid19 on their lives.

## 2 Methodology

The present analysis is inspired on Cross Industry Standard Process for Data Mining(CRISP-DM)^35^ steps, the phases are very frequent on Data Mining tasks. So, the steps for this analysis are the next:

- Load dataset from collection
- Choose relevant terms associated to the problem
- Select posts
- Cleaning data to eliminate words with no relevance(stopwords)
- Visualization to support on the understanding

### 2.1 Loading dataset

There is a limitation when the collection of posts is performed so the query only can get seven past days and for this reason is necessary to collect weekly according to the problem to analyze. On this context, the data collect from Bogota is useful to analyze the impact of covid19 on Mental Health of population living in Bogota, Colombia.

### 2.2 Choose relevant terms

The previous terms selected to collect posts were covid19, coronavirus and some derivations using ‘-’,’#’, ‘@’, i.e coronavirus, @covid19 then publications related to covid19 were selected. For the actual context, terms related to anxiety and some psychological effects coming from lockdown and people at home are the next:

- miedo, indiferencia (indiferente), morir
- familiar, sobrevivir, infectar, salud mental, estré
- seres queridos, familia,violencia ins, ansiedad, rabia, solo
- soledad, duelo, depresión, salir, aire, abrumado, abrumada
- resiliencia, dieta alcalina

### 2.3 Cleaning data

- Uppercase to lowercase
- Eliminate special characters
- Eliminate stopwords
- Eliminate words with size less than 3

### 2.4 Visualization

For the understanding of the impact on the population,…

## 3 Results

The next graphics presents the results of the experiments and answer many questions to understand the phenomenon over the population.

### 3.1 How frequent are the publications related to covid19 and Mental Health

At the beginning of the pandemic in Bogota, Colombia there was a few posts, a open question is: was there some previous problems related to the scope before covid19? Considering figure Fig.7, the evidence presents the increasing number of post during the next days and a peak during the range of March 16th - 21th.

**Figure 1.**
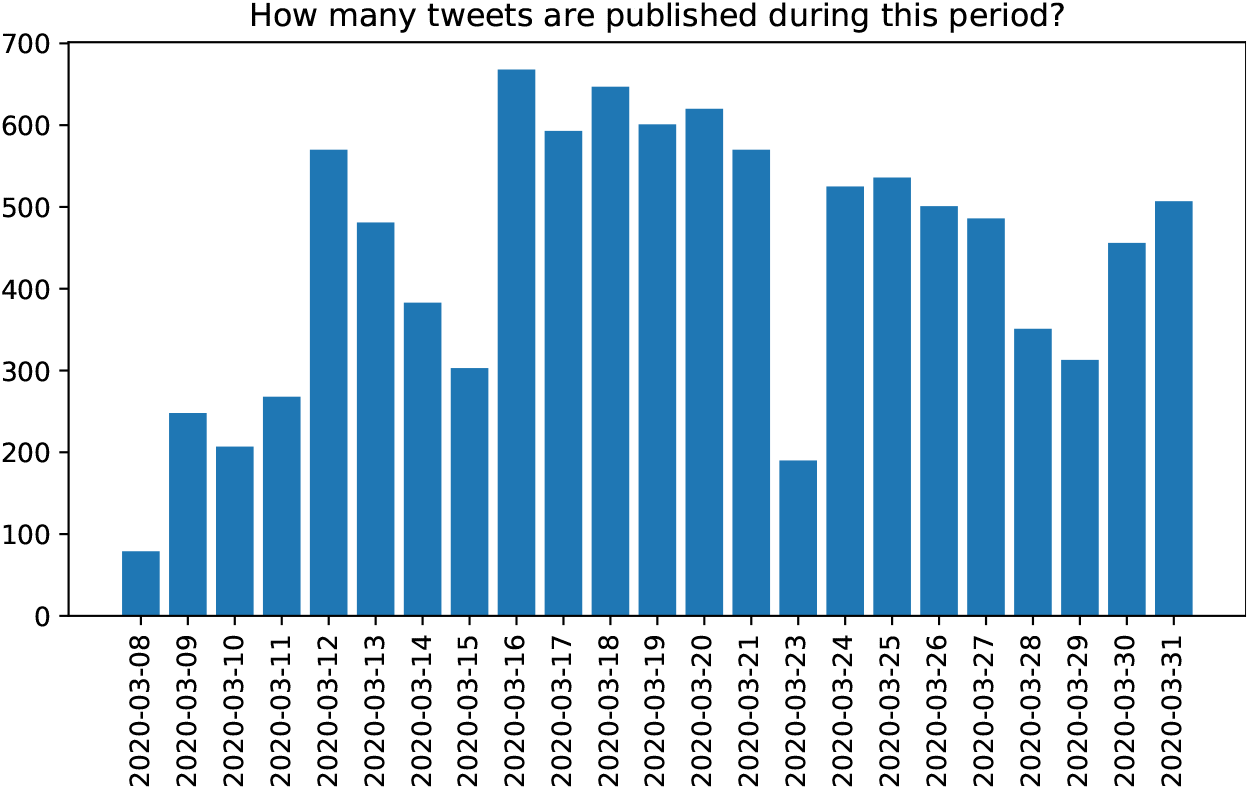
Data User Creation.

**Figure 2.**
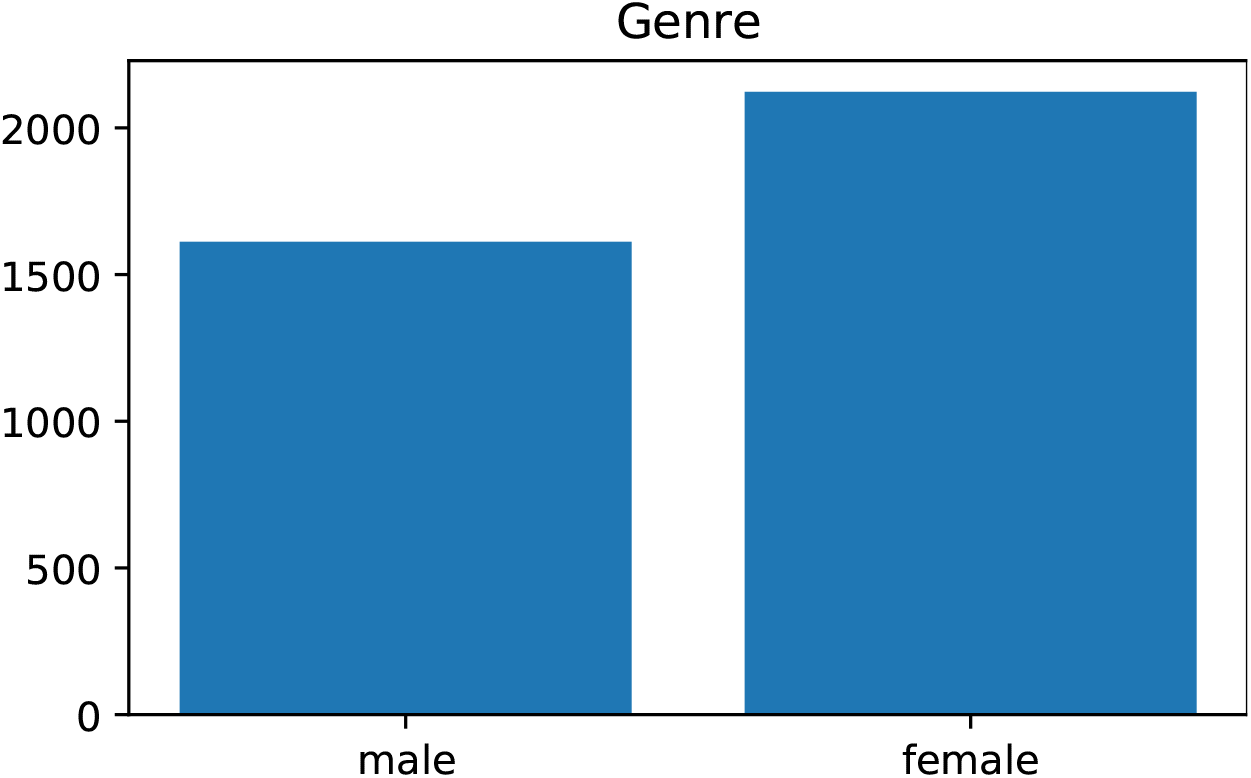
Data User Creation.

**Figure 3.**
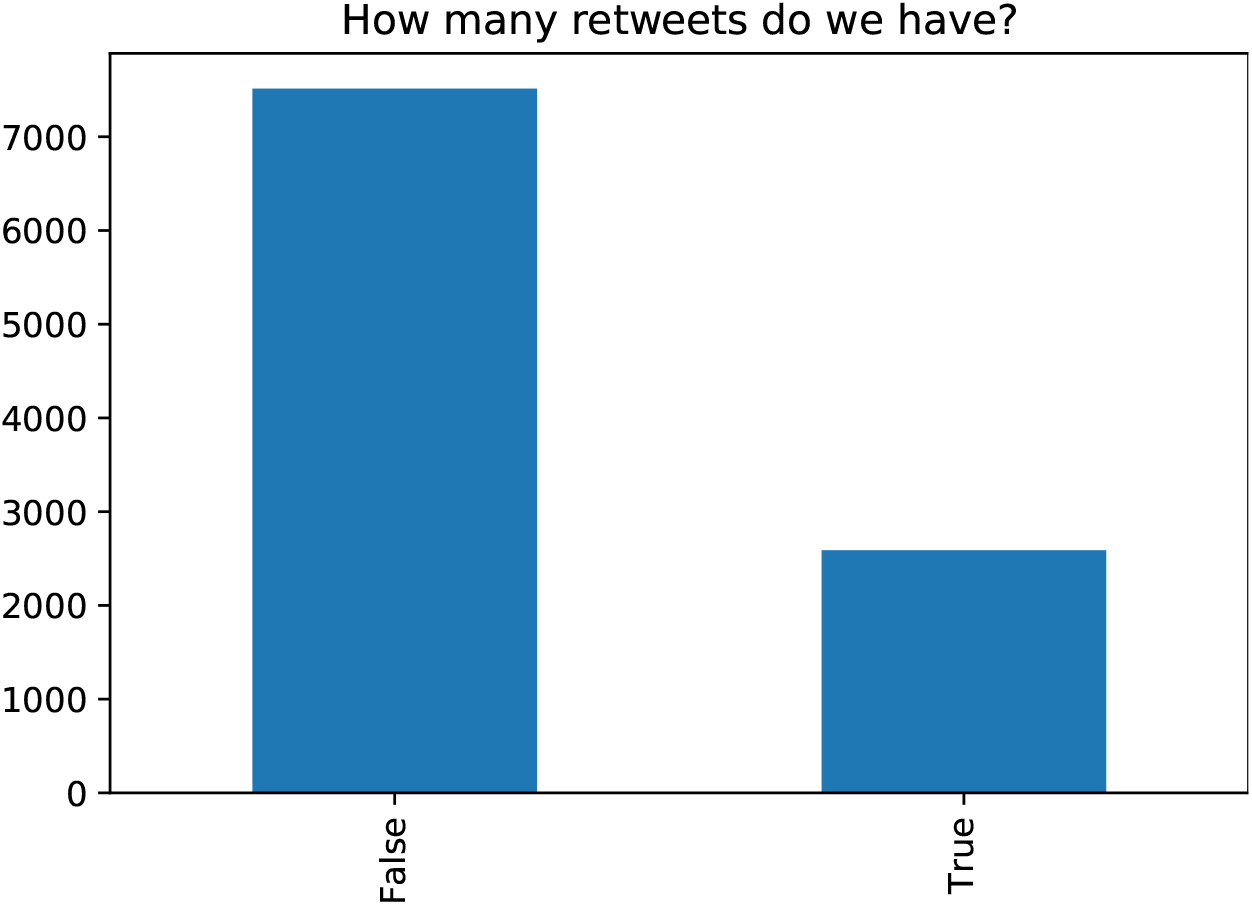
Data User Creation.

**Figure 4.**
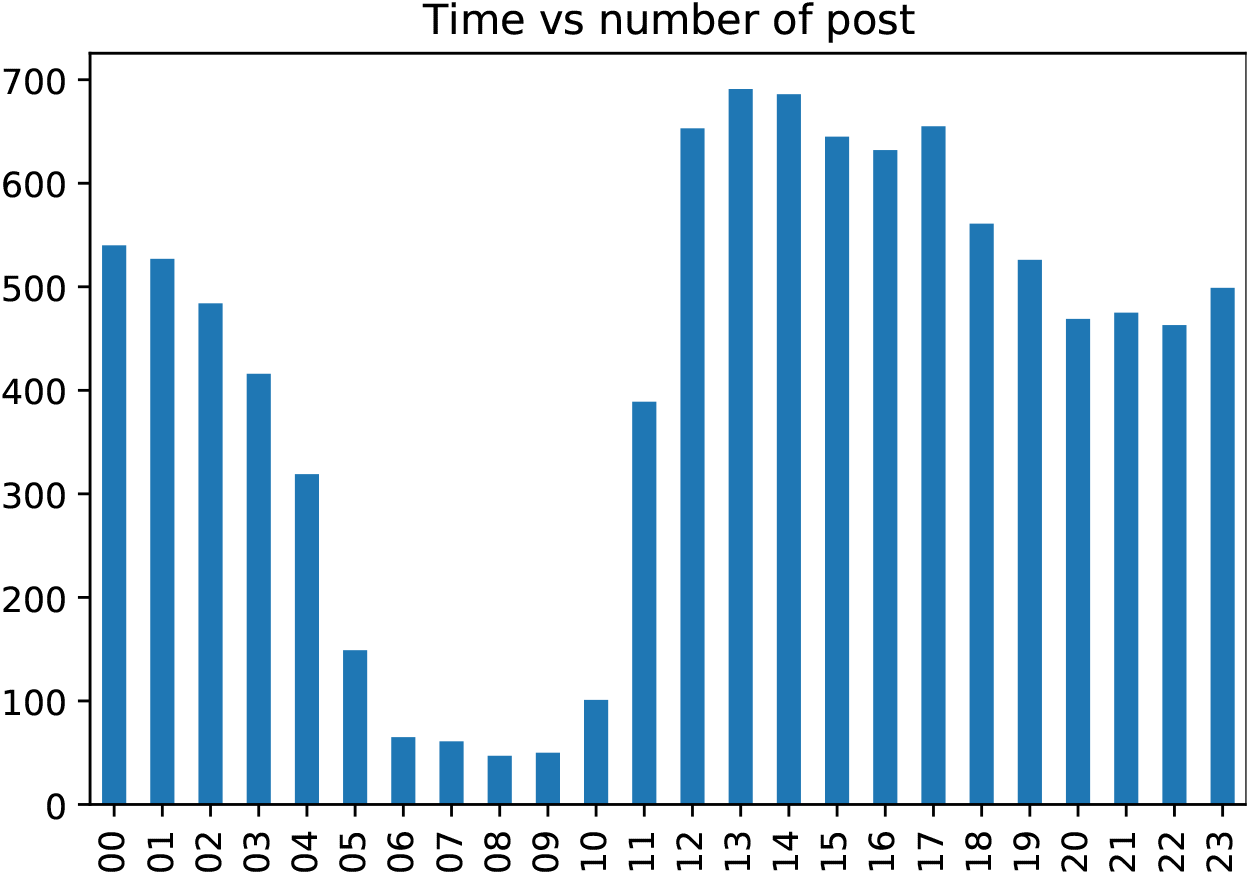
Data User Creation.

**Figure 5.**
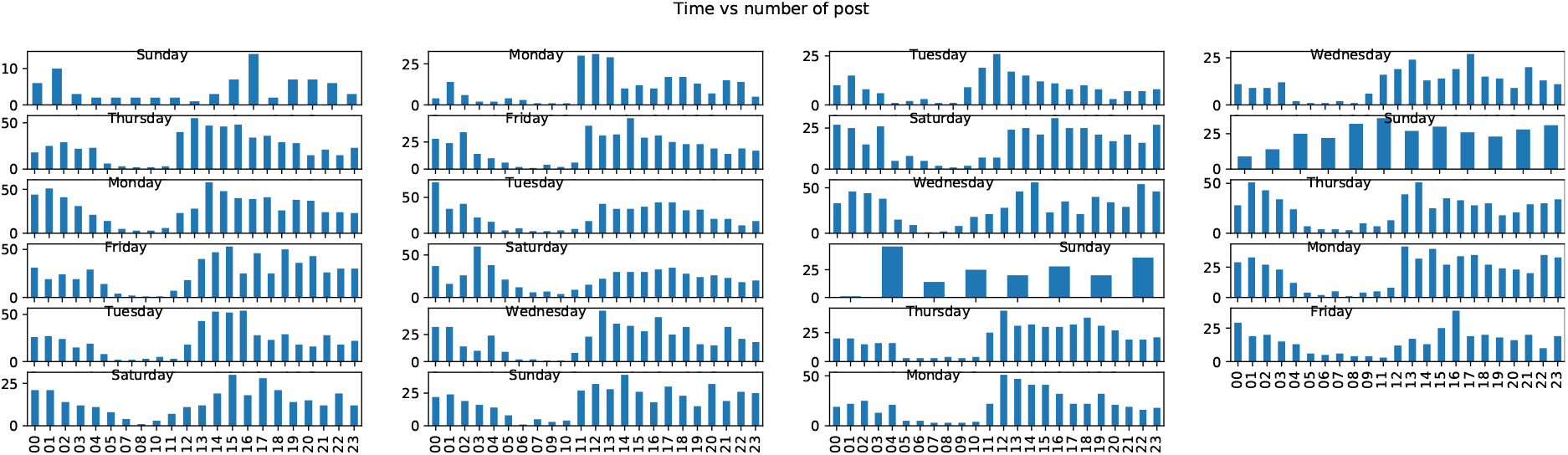
Data User Creation.

**Figure 6.**
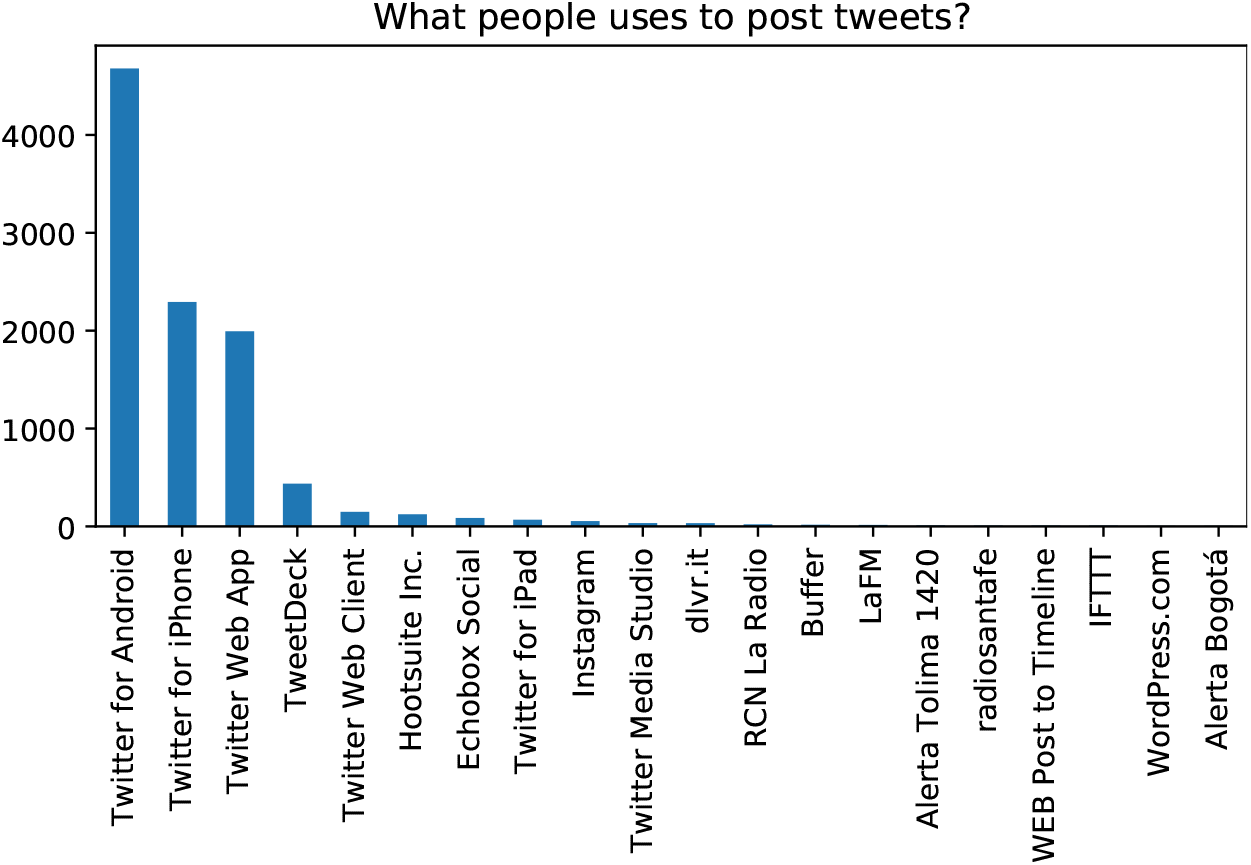
Data User Creation.

**Figure 7.**
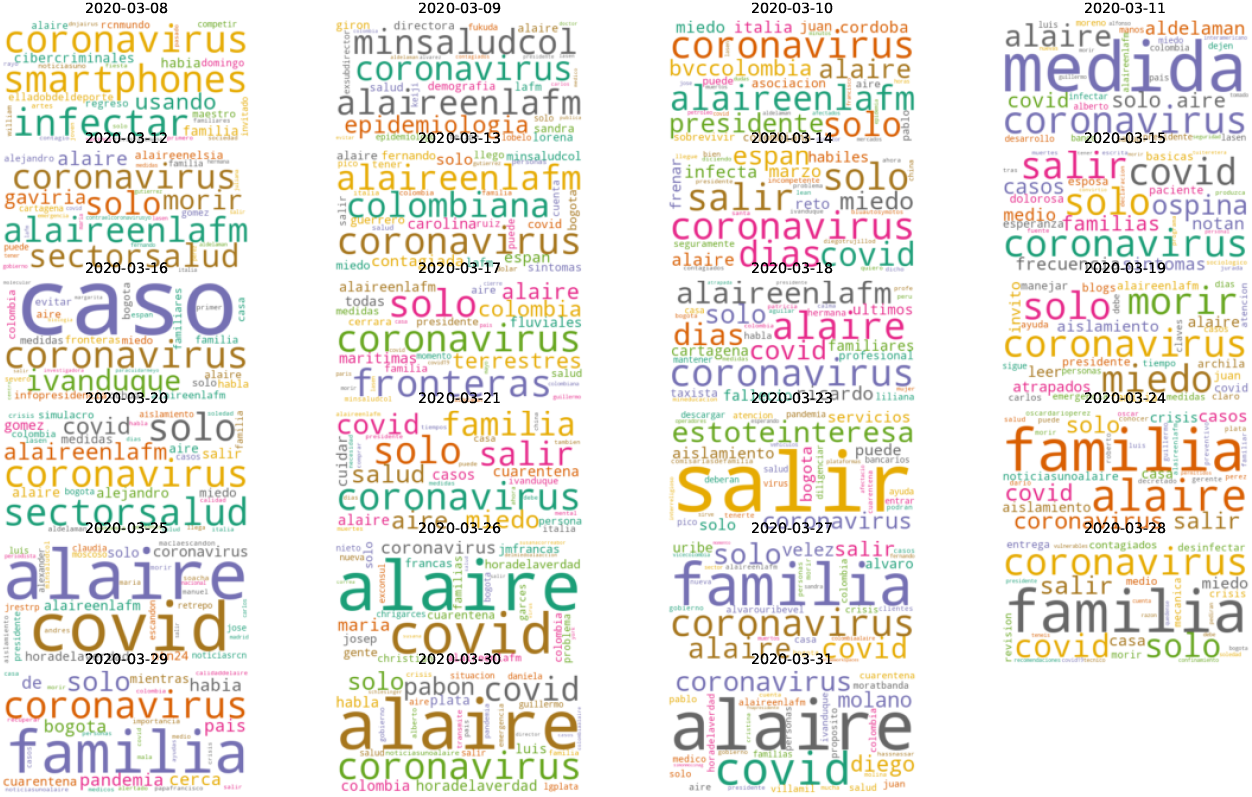
Data User Creation.

### 3.2 How is the distribution of women/men tweets

Actually, any information about genre is found in tweets but there is a artificial solution to know about it. Considering the next criterion: usually women names have a termination with letter ‘a’ and men have an ‘o’ at the end. Then, the graphic Fig.2 and the next affirmation is possible, women are posting more about how they are feeling facing covid19 isolation at home.

### 3.3 People share their own thoughts

On Twitter, if one person likes or agrees with some content, the retweet occurs to share what users want. One question related to this is: people writes and shares about their own concerns or prefer to talk through others. The bar plot Fig.3 explains us Colombian users prefers to write their own content.

### 3.4 Post distribution per hour

Lockdown start on March, so many people went to home for home office or got fired during this period. More time at home means face daily issues with family for more time or long periods than ever. Usually, employees work from 8-18h so there was natural to have a increase number of posts after work and a meaningful decreasing number after 23h but graphic Fig. 4 are showing other behaviour, this pattern can be related to disorder dreams.

And what is happening during the days, users are online all the days and post at the same time everyday. The illustration 5 of the hour frequency post explains a common patter during the morning and night.

### 3.5 What kind of device

There is many ways to connect and use Twitter SN, i.e. laptops, cellphones, tablets, etc. The most common device is cellphone and those have Android and Ios, from the graphic Fig. 6 is evident Android is more common and people preferences to use cellphones to be online. This graphic could be usefutl to corelate social information with kind of device, using the relation: ‘people with good income can afford Iphones and the opposite: ‘people with short saleries can pay for Androids.

### 3.6 Frequent term during the weeks

Terms and bigrams presented in figures 7 and 8 confirmed some finding in the work of Bonrud and Ahorsu about fear, anxiety, isolation, stress. All this features can influence on the population, considering sleep habits can notice how they are changing. Besides, colombian families have family outside of Colombia, i.e. tourism during the evolution of covid19 in Colombia, people started to worry about how they are going to come back.

**Figure 8.**
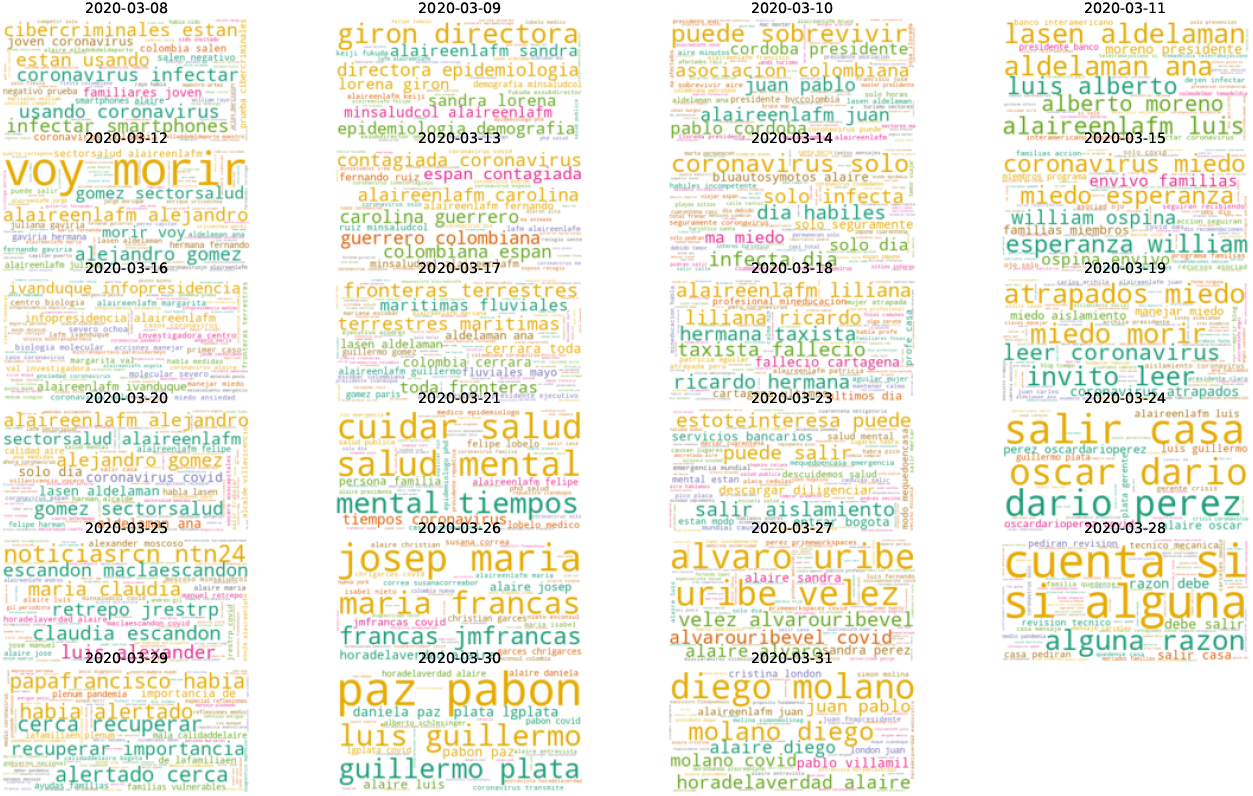
Data User Creation.

## 4 Conclusions

Infoveillance based on Social Sensors with data coming from Twitter can help to understand the trends on the population of the capitals. In this context, analyze what people posted during the month of March can help to understand the behaviour of people and know what kind of disorders or issues they faced to promote better actions to help them.

## Data Availability

The data can be accesible asking it.

